# Antibody cross-reactivity and evidence of susceptibility to emerging Flaviviruses in the dengue-endemic Brazilian Amazon

**DOI:** 10.1101/2022.06.07.22276118

**Authors:** Barbara Batista Salgado, Fábio Carmona de Jesus Maués, Maele Jordão, Renato Lemos Pereira, Daniel A. Toledo-Teixeira, Pierina L. Parise, Fabiana Granja, Higo Fernando Santos Souza, Marcio Massao Yamamoto, Jannifer Oliveira Chiang, Livia Caricio Martins, Silvia Beatriz Boscardin, Jaila Dias Borges Lalwani, Pedro Fernando C Vasconcelos, José Luiz Proença-Modena, Pritesh Lalwani

## Abstract

Flaviviruses are vector-borne positive sense RNA viruses with enormous potential to cause a spectrum of severe diseases in naïve populations and long-term public health impact. Several of these Flavivirus es can co-circulate; recently, Zika virus (ZIKV) is becoming increasingly prevalent in Dengue virus (DENV) endemic regions. Pre-existing immunity to one virus can modulate the response to a heterologous virus; however, the serological cross-reaction between these emerging viruses in DENV endemic region are poorly understood. In this study, we estimated the seropositivity rates of nine different Flavivirus es among residents of Manaus city in Brazil. Next, we assessed antibody cross-reactivity of DENV-positive individuals with ZIKV using hemagglutination inhibition assay (HIA), purified envelope proteins in an ELISA, and live virus neutralization test. 74.52% of participants were IgG positive (310/416) as estimated by lateral flow tests. Overall, 93.7% of participants were seropositive (419/447) for at least one DENV serotype and the DENV seropositivity ranged between 84.8% and 91.0% as determined by HIA. DENV positive individuals with high antibody titers in HIA or envelope protein domain III (EDIII)-ELISA reacted strongly with ZIKV, whereas individuals with low anti-DENV antibody titers reacted poorly towards ZIKV. Live virus neutralization test with ZIKV confirmed that dengue serogroup and ZIKV-spondweni serogroup are distant and DENV-positive individuals do not cross-neutralize ZIKV efficiently. Taken together, we observed a high prevalence of DENV in the Manaus-Amazon region and a varying degree of serological reactivity against emerging viruses. Ecological and epidemiological conditions in Manaus makes its population susceptible for further arbovirus outbreaks; hence functional vector control program and febrile-syndrome surveillance are essential to identify unforeseen epidemiological threats.

## Introduction

Emerging arboviruses are a growing health problem in Brazil and worldwide. During the last decade, Brazil has endured zika virus (ZIKV), chikungunya virus (CHIKV), and yellow fever virus (YFV) epidemics. These pathogens have not only been associated with morbidity and mortality, but also with additional health care cost. Arboviruses such as DENV and YFV have been endemic in Brazil, while YFV is endemic/enzootic for centuries being introduced during the slave traffic, DENV is more recently in the country and endemic just to four decades. Recent studies have demonstrated an increase in dengue-associated deaths in the last decade. Yellow fever vaccine has been shown to be efficient in preventing infections, however, recent YFV epidemic in Southeast Brazil demands better understanding on the transmission dynamics and disease distribution in general population [1].

The introduction of ZIKV in Brazil caused a huge epidemic in some regions of Brazil, which was associated with microcephaly in the newborns [2, 3] and Guillain-Barré syndrome and few fatal cases in adults [4, 5]. New studies have shed light on the ever-growing pathologies associated with ZIKV infection. A high density of *Aedes* mosquitoes in several cities was responsible for the high transmission rates; however, still little is understood about the interaction between DENV and ZIKV and the underlying cause of the magnitude of the ZIKV epidemic in Brazil.

A high proportion of dengue infections produce no symptoms or minimal symptoms, especially in children and in those with no previous history of dengue infection. Prior DENV infection increases the risk of future symptomatic and Dengue Hemorrhagic Fever (DHF), a severe and sometimes fatal clinical disease presentation. Non-neutralizing antibodies against the envelope protein induced during primary DENV infection can cross-react with other DENV serotypes or heterologous Flavivirus es. These DENV cross-reactive antibodies can facilitate subsequent secondary or tertiary heterologous DENV infection of myeloid cells due the antibody-dependent enhancement (ADE) and can increase dengue disease severity in humans [6]. Similarly, DENV serotypes and other Flavivirus es share peptides that can modulate protection and the other aggravate the pathogenesis by CD4+ and CD8+T cell responses [7].

*In vitro* and mouse challenge studies have shown that antibodies raised against DENV can enhance ZIKV infection [8, 9]. However, prior DENV infection was not associated with ZIKV viremia or cytokine expression in experimentally challenged macaques [10-12] or in humans [13-15], nor with fetal demise or congenital Zika syndrome in pregnant women [16, 17]. Emerging evidence suggests that prior DENV infection may not enhance non-congenital Zika disease, but whether prior ZIKV infection increases future dengue disease in humans remains unknown. Nevertheless, dengue endemic regions with YFV vaccination could have partially played a role in protecting individuals against ZIKV severe disease rather than participating in increasing ZIKV replication due to the presence of antigenically related immune memory. In this study, we estimate seropositivity rates against nine different endemic or emerging *Flavivirus* genus members in Manaus, capital of the Amazonas state. Next, we assessed antibody cross-reactivity of DENV positive individuals with ZIKV using hemagglutination inhibition assay (HIA), purified envelope proteins in an ELISA and live virus neutralization test.

## Methods

### Ethics

This observational and cross-sectional study was approved by the Research Ethics Committee of the Universidade Federal do Amazonas (UFAM), with approval number CAAE 96171218.7.0000.5020, in accordance with the Brazilian law, which complied with the Declaration of Helsinki. All study participants gave oral and written informed consent prior to enrollment.

### Study population and sample and data collection

The study population comprised of healthy individuals accessing services at the Centro de Controle de Zoonoses (CCZ), Manaus-Amazonas. Four hundred and fifty consecutive participants were recruited between January 2015 and December 2015. The study included individuals of both sexes ≥18 years old who agreed to participate. Study participants signed the consent form and answered an epidemiological questionnaire. Four mL of venous blood was drawn from each participant using EDTA tubes (BD Vacutainer). Soon after blood collection, collection tubes were centrifuged, and plasma was separated and stored at -80°C until further analysis.

### Sample size calculation

A sample size of 398 individuals was calculated using an estimated prevalence of 50% and 95% confidence interval for a large population with a desired precision of 0.01 [18]. To avoid dropouts due to incomplete questionnaires and subsequent reduction in statistical power, we recruited 450 individuals. However, three questionnaires were incomplete and removed from the analysis. Sample size calculation was performed using the Epi Info software version 7.2.

### Maps and socioeconomic indicators from Brazil

Maps were created using the QGIS Software version 2.18.26 for macOS. Graphs displaying the Human Development Indexes and Sanitation Indicators using public data available in Atlas Brasil from the 2010 Census (https://atlasbrasil.org.br) and the National System of Sanitation Information/Trata Brasil from 2018 (http://www.snis.gov.br), respectively.

### Dengue and Zika cases in Manaus and Brazil

Supplementary Figure 1 compares DENV and ZIKV cases before and after the Zika epidemic in Manaus city and Brazil. Graphs were plotted using Dengue and Zika confirmed cases by epidemiological week and year of first symptoms, between 2015 and 2017. These data are changeable and were available on Tabnet–DATASUS (Ministry of Health Brazil, https://datasus.saude.gov.br/informacoes-de-saude-tabnet/), and PLISA– PAHO/WHO (Health Information Platform for the Americas –Health Pan-American Organization, https://www3.paho.org/data/index.php/es/).

### Dengue Rapid Test

Lateral flow Point of Care (POC) Dengue virus antibodies detection test was performed using the BioPix® Dengue IgM/IgG Rapid test (Wama Diagnóstica, São Paulo, Brazil) at the Laboratory of Emerging Viruses at the University of Campinas (LEVE - Unicamp). The plasma samples were assayed to qualitatively detect IgG and IgM antibodies against all four Dengue virus serotypes. The assay had a sensitivity of 99% and specificity of 98% as declared by the manufacturer. The detection of IgG in this assay was developed to detect low levels of IgG.

### Hemagglutination inhibition test

To assess overall Flavivirus reactivity, HIA was performed at Evandro Chagas Institute (IEC), Belém. Serum samples collected were subjected to a hemagglutination inhibition assay (HIA) and adapted to the microplate technique, with a titration cut-off point of 1/20 as previously described. Plasma samples were tested for antibodies against the yellow fever vaccine strain (YFV-17D) and 10 endemic or emerging arboviruses of the Flavivirus genus in the Brazilian Amazon: Yellow Fever virus (YFV), Dengue Virus (DENV) serotypes 1 to 4 (DENV-1, DENV-2, DENV-3 and DENV-4), Zika virus (ZIKV), Saint Louis Encephalitis virus (SLEV), West Nile virus (WNV), Ilheus virus (ILHV), and Rocio virus (ROCV)].

For the HIA test, plasma samples were pretreated with acetone (P.A.) and NaCl (0.85%) solution, hydrated with borate buffer saline (BBS) and adsorbed with goose (*Anser cinereus*) erythrocytes. In the screening step, they were added to a 96-well microtiter plate (‘U’ shape wells) together with a viral suspension containing four hemagglutinating units (4 HAU) of the test antigen and a suspension of white goose erythrocytes diluted in dextrose, gelatin, and barbital at 1:5 dilution (DGV). Initially, the screening was performed at 1:20 dilution, then positive samples were serially diluted with bovine serum albumin (0.4%) up to a 1:1280 dilution.

### Cloning, expression, and purification of recombinant DENV2 and ZIKV envelope domain III proteins

The DENV2 envelope domain III protein sequence (E-DIII, residues 577-674, GenBank number HQ026763) was cloned and expressed using the pET28b vector [Silvia Beatriz Boscardin, University of São Paulo (USP)] as previously described [19]. Zika virus envelope amino acid sequences from different isolates was obtained from GenBank and multiple sequence alignments was performed to identify an amino acid consensus sequence using MEGA software version 7.0.26. A polyhistidine (6x-His) tag was added at the N-terminal to facilitate downstream purification. Consensus ZIKV E-DIII sequence was codon-optimized and inserted into prokaryotic expression vector pGS21-a (GenOne Biotechnologies, Brazil). Recombinant vectors were transformed into electrocompetent Escherichia coli BL21 (DE3) cells, after incubation ampicillin-resistant colonies were grown at an optical density of 0.6-0.8 (600nm) at 30°C. Protein expression was induced with 1 mM isopropylD-1-thiogalactopyranoside (IPTG). The bacterial culture was harvested by centrifugation at 4,000 rpm for 20 minutes at 4°C. Pellets were resuspended in lysis buffer (lysozyme 1mg/mL and PMSF 100 mM) and lysed by ultrasonication; and debris were removed by centrifugation. The remaining sediments were subjected to another round of lysis with lysis buffer, sonication, and centrifugation. The bacterial lysate was resuspended in denaturing extraction buffer (NaH2PO4100mM, Tris-HCl 10 mM, Urea 8M, pH 7,0), and debris were separated by centrifugation. Post-extraction supernatants containing E-DIII proteins were purified by metal affinity chromatography using a Ni-NTA column (HisTrap® HP column 1mL, GEHealthcare Life Science) coupled to an ÄKTA Purifier 10 system (GE healthcare) using a gradient; proteins were eluted in buffer containing NaH2PO4 100mM, Tris-HCl 20 mM, Urea 8M, 500 mM imidazole, glycerol 10%, pH 7.0. Affinity purified proteins were then applied to Amicon® Ultra-2mL 3K Centrifugal Filters (MerckMillipore®) for desalting, diafiltration, concentration, and buffer exchange to phosphate buffer saline (PBS) pH 7.4, following the manufacturer’s recommendations.

### SDS-PAGE and Western Blot analysis

The soluble fractions of purified DENV- or ZIKV-E-DIII proteins collected during the purification process were analyzed on an SDS-PAGE stained with CoomassieBlue. Western blotting (WB) was used to test the antigenicity of the recombinant proteins (Supplementary Figure 3). DENV or ZIKV-EDIII proteins were loaded onto 12% polyacrylamide gels and transferred to a nitrocellulose membrane (0.2uM hybond ECL – GE Healthcare) by semi-dry electroblotting method. Transferred membranes were blocked with 5% skim milk in PBS for one hour at room temperature. Membranes were then incubated for one hour at room temperature with mouse anti-His tag antibody (Sigma), or DENV or ZIKV positive and negative human samples at a 1:1000 dilution. After incubation, membranes were washed three times with PBS-Tween 20 0.05% (PBS-T). Next, secondary anti-mouse or anti-human horseradish labeled IgG antibodies were added at 1:1000 (KPL, Seracare, USA) and incubated for one hour at room temperature. After incubation, excess unbound antibodies were removed by washing the membrane three times with PBS-T. The nitrocellulose membranes were then incubated with DAB (3,3’,4,4’ diaminobenzidine) (BioRad, EUA) plus hydrogen peroxide for 5 minutes, the reaction was stopped by washing the membranes with distilled water. Membranes were air dried and protein bands were then compared to the protein molecular weight marker (Sigma).

### ELISA to detect anti-DENV and ZIKV E-DIII-specific IgG antibodies

We standardized an indirect in-house Enzyme Linked Immunosorbent Assay (ELISA) to detect anti-DENV and anti-ZIKV IgG antibodies in plasma samples using recombinant E-DIII proteins as antigens. The checkerboard method was used to determine antigen concentration, sample dilution, and secondary antibody concentration to obtain an optimal signal-to-noise ratio. To test patient samples, 96-well ELISA plates (Immunoplate, SPL) were coated with 100 ng of DENV or ZIKV E-DIII protein in carbonate buffer (0.015 M Na2CO3, 0.035 M NaHCO3 pH 9.6) at 4oC overnight and unbound proteins were removed by washing 5 times with PBS-T (0.5% Tween). Wells were then blocked with 10% non-fat dry milk in PBS for 90 minutes at room temperature and orbital shaking and plates were then washed 4 times with PBS-T. Plasma samples at a 1:100 dilution were incubated at room temperature for 1 hour, followed by washing 5 times with PBS-T. Next, an anti-human horseradish peroxidase-labeled IgG antibody produced in goat (KP, Seracare, USA) was added at a 1:10,000 concentration to each well, plates were incubated at room temperature and then washed 5 times with PBS-T. TMB (3,3′,5,5′-Tetramethylbenzidine) substrate (Thermo Fisher Scientific) was added to the antigen-antibody enzyme complex and incubated for 5 minutes at room temperature. The reaction was stopped by adding 3M of sulfuric acid. The absorbance optical density was read at 450 nm with a microplate reader (Hidex chameleon V microplate reader).

### Zika virus neutralization test

Focus reduction neutralization test (FRNT) was performed in collaboration withthe Laboratory of Emerging Viruses at University of Campinas (LEVE – Unicamp). Serum samples were heated at 56°C for 30 minutes to inactivate the complement system. 4-fold plasma samples diluted with Earle minimum essential medium (MEM) (Sigma-Aldrich) were incubated with 100 focus forming units (FFU) of ZIKV (strain BeH823339-Asian). This virus plasma mixture was then added to a monolayer of Vero-E6 cells (ATCC) cultivated in 96-well plates (Corning) and maintained at 37°C with CO2 5% for 2 hours. Semi-solid media (MEM, medium viscosity carboxymethyl cellulose 0.75%, fetal bovine serum 5%, and penicillin and streptomycin 1%) was added to each well and plates incubated for another 48 hours at 37°C with 5% CO2. 1% paraformaldehyde was added to the 96 well plates and fixed overnight at 4°C. Next, cells were washed 3 times with PBS pH 7.4, blocked and permeabilized by adding washing buffer (PBS 0.15M, bovine albumin serum 0.1%, Triton X-100 0.1%) for 30 minutes. Next, 1:1,000 anti-ZIKV NS1 antibody (Abcam) was added, and plates were incubated for 2 hours at room temperature (20 to 25°C). Cells were subsequently washed and incubated with 1:2,000 anti-mouse IgG peroxidase-conjugated (Sigma-Aldrich) for 1 hour at room temperature. Cells were then washed and TrueBlue peroxidase substrate (KPL) was added following the manufacturer’s recommendations. FFU units were then counted for each plasma dilution and the results were tabulated.

### Statistical Analysis

Descriptive statistics was used to describe the socio-demographic features and mosquito preventive measures of the study population. Missing values were excluded from the calculations. Bubble charts showing the percentages of seropositives with HIA antibody titers were created with Microsoft Excel 2019 software. 95% CIs were computed by Blaker’s method. All statistical analyzes were performed using the Graphpad Prism software version 9.1.2. Paired T-test was used to evaluate the ELISA results for DENV- and ZIKV-E-DIII. The linear correlation between variables was determined using Pearson’s correlation. P values ≤0.05 were considered statistically significant.

## Results

### High dengue prevalence in urban Manaus

A random convenience sampling strategy enrolled 450 individuals in Manaus, this noninterventional cross-sectional study between January and December 2015 (Supplementary Figure 1). 447 healthy individuals of both sexes and ≥18 years were included in the final analysis; 3 questionnaires were incomplete and removed from the analysis. Demographic features of the study population are described in Table 1. We oversampled females (70.5%) and the median age of our study population was 38 years. 30% of the study participants auto declared that they were unemployed in the last 12 months and 61.70% had completed high school (Table 1). 37.5% and 13.2% of the study participants self-reported a previous dengue and malaria infection, respectively. 80.8% reported vaccination for YFV and 75.9% use SUS-Brazil’s public health system. A majority (94.9%) reported performing at least one preventive measure against mosquitoes: avoiding standing water and properly disposing of trash, followed by personal mosquito repellents, were the most popular measures adopted to prevent mosquitoes (Table 2).

**Table 1.**
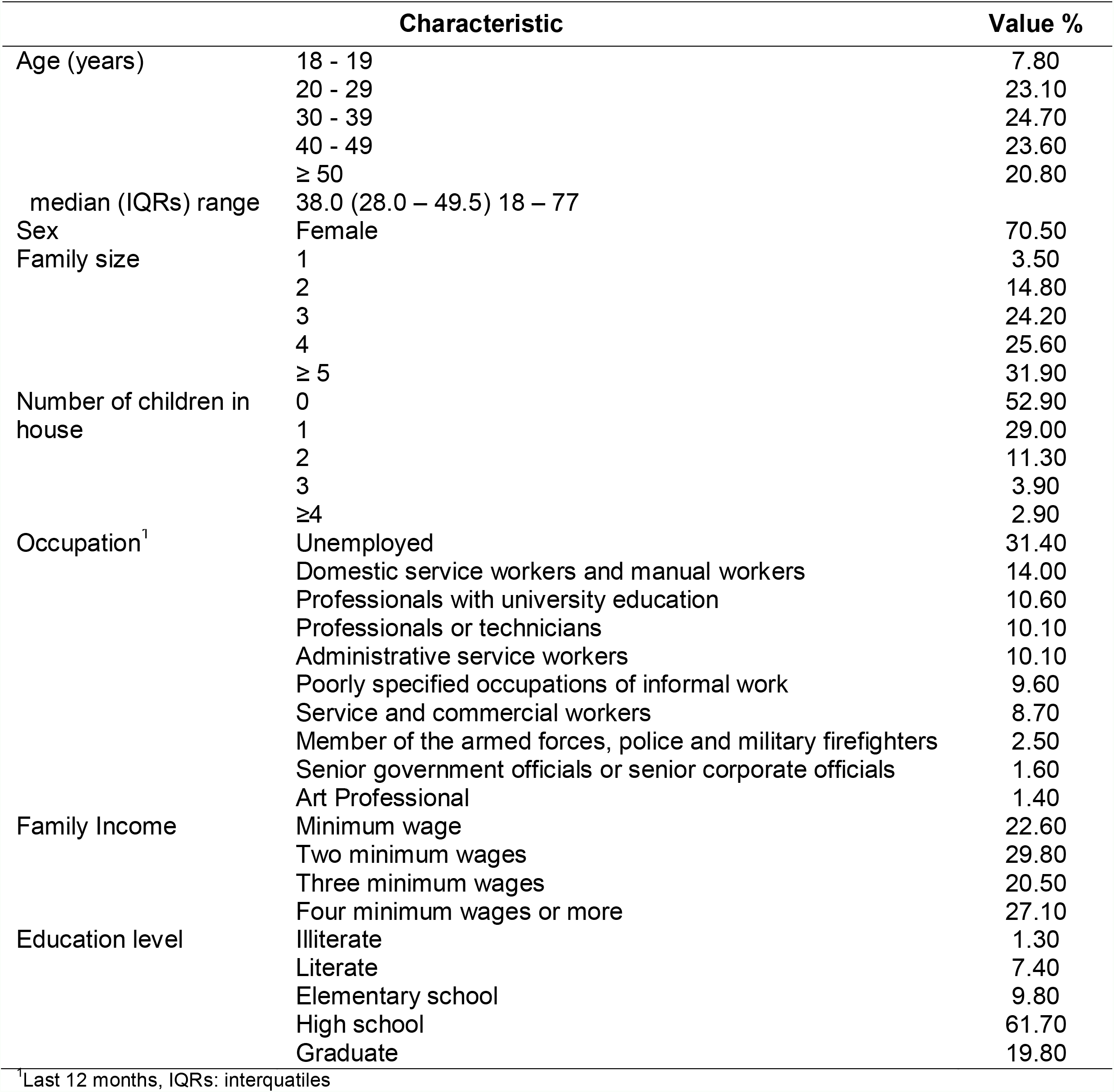
Sociodemographic features of the study population-Manaus, Amazonas

**Table 2.** Mosquito prevention practices reported by study participants

| Characteristic |  | Value % |
| --- | --- | --- |
| Dengue infection <sup>1</sup> | Yes | 37.5 |
| Malaria infection <sup>1</sup> | Yes | 13.2 |
| Vaccinated against Yellow fever <sup>1</sup> | Yes | 80.8 |
| Use Sistema Único de Saúde (SUS) <sup>2</sup> | Yes | 75.9 |
| Neighbours with dengue fever | Yes | 43.3 |
| Mosquito problem in neighbourhood | Yes | 43.1 |
| Perform preventive measures against mosquitos | Yes | 94.9 |
| Mosquito prevention measures <sup>3</sup>: |  |  |
|  | Avoid standing water | 64.30 |
|  | Disposing trash correctly | 62.50 |
|  | Personal mosquito repellents: cream and spray | 54.90 |
|  | Personal and household hygiene | 33.70 |
|  | Fans | 28.10 |
|  | Close windows and doors | 21.80 |
|  | Mosquito coil, mats and liquid vaporisers | 12.60 |
|  | Mosquito net for door and window | 10.90 |
|  | Protective clothing | 9.40 |
|  | Mosquito bed nets | 5.80 |
|  | Others | 4.30 |

<sup>1</sup> Self-reported, <sup>2</sup> Brazilian public funded health care system, <sup>3</sup> Multiple responses

Manaus is a tropical metropolis in Brazil with about 2.5 million inhabitants in the middle of the Amazon rainforest (Figure 1A). Manaus, the capital of the Amazonas state, compared to other Brazilian capitals, shows a low sociodemographic and human development index (HDI). In Manaus, the lack of sanitation is also widespread, with wastewater facilitating vector proliferation and waterborne diseases, which affect the poorest (Figure 1B). Figure 1C compares our study population age distribution compared to the 2010 census data.

**Figure 1.**
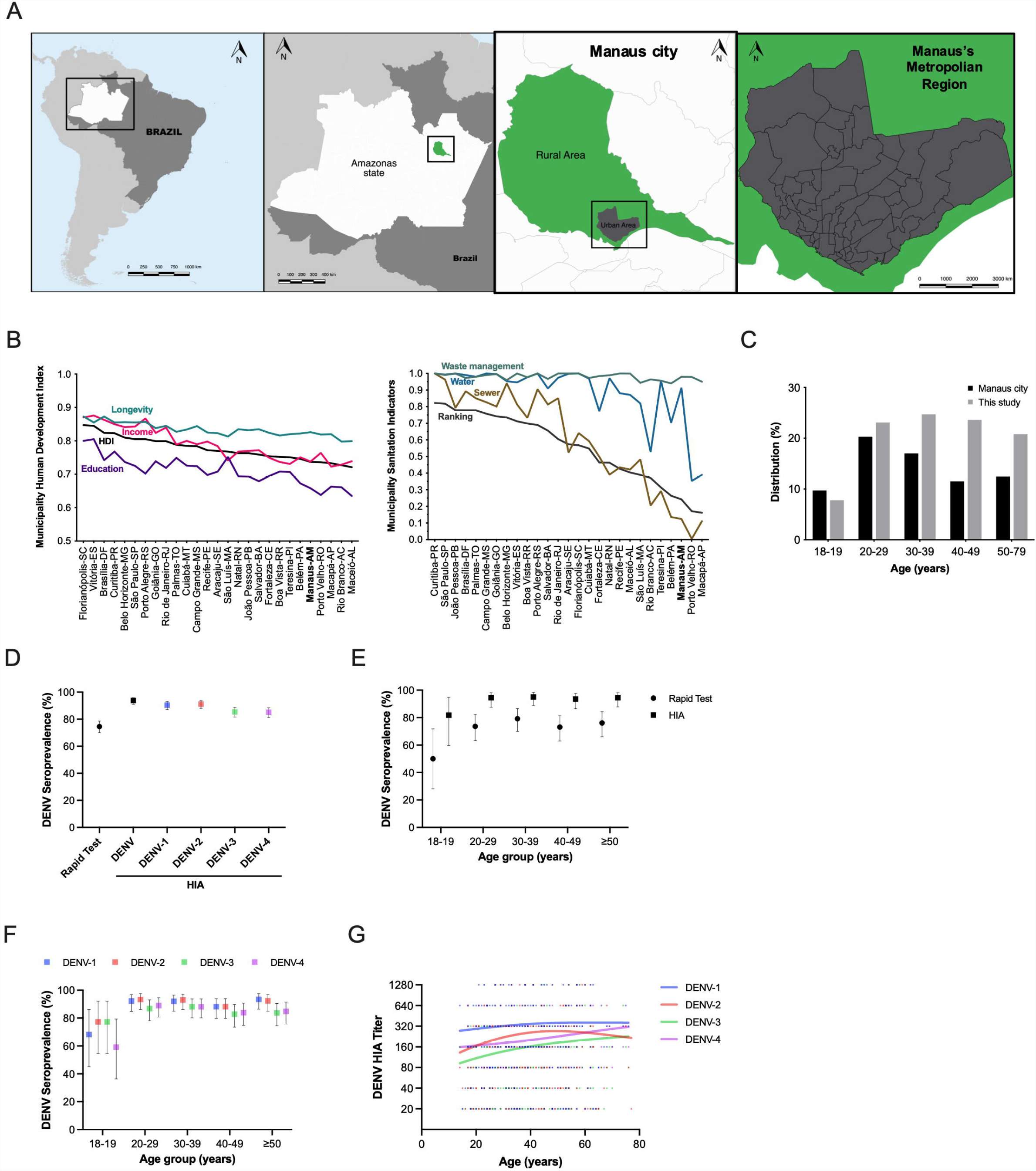
Elevated Dengue seropositivity rates in Urban Manaus (A) Maps depict location of Manaus. (B) Human Development Index (left) and Sanitation indicators (right) for the 27 Brazilian capitals were obtained from the 2010 census and a 2018 survey, respectively. (C) The study population distribution is compared to 2010 Census data obtained from the Brazilian Institute of Geography and Statistics (IBGE). Plasma samples were tested for anti-Dengue virus antibodies using (D-E) BioPix® Dengue Rapid Test IgG/IgM (n=416) and (D-G) Hemagglutination Inhibition Assay (HIA, n=447) and seropositivity percentages with 95%CI were plotted. BioPix® Dengue Rapid Test IgG/IgM was qualitative. HIA was quantitative and samples with HIA titer ≥20 units were considered positive for the test virus.

Next, we assessed the prevalence of anti-DENV antibodies using lateral flow point of care (POC) rapid tests and hemagglutination inhibition assay (HIA). 74.52% of participants were IgG positive (310/416) and 1.68% (15/416) had anti-DENV IgM antibodies as estimated by POC tests. Overall, 93.7% of participants were seropositive (419/447) for at least one DENV serotype and the DENV seropositivity ranged between 84.8 and 91.0% as determined by HIA (Figure 1D and Table 3). We observed that most of our participants over 20 years of age had anti-DENV antibodies (Figure 1E-G and supplementary Figure 2).

**Table 3.** Prevalence of anti-Flavivirus antibodies

| Serogroup | Virus* | Total# % | Age (years) |  |  |  |  |
| --- | --- | --- | --- | --- | --- | --- | --- |
|  |  |  | 18-19 | 20 - 29 | 30 - 39 | 40 - 49 | ≥ 50 |
| Yellow Fever Vaccine | YFV/17D | 92.8 (415/447) | 67.6 (23/34) | 98.0 (99/101) | 95.4 (103/108) | 93.2 (96/103) | 92.3 (84/91) |
| Yellow Fever Virus Group | YFV | 80.5 (360/447) | 61.8 (21/34) | 77.2 (78/101) | 83.3 (90/108) | 79.6 (82/103) | 86.8 (79/91) |
| Dengue Virus Group | DENV® | 93.7 (419/447) | 79.4 (27/34) | 95.0 (96/101) | 95.4 (103/108) | 94.2 (97/103) | 95.6 (87/91) |
|  | DENV-1 | 90.6 (405/447) | 76.5 (26/34) | 93.1 (94/101) | 91.7 (99/108) | 88.3 (91/103) | 93.4 (85/91) |
|  | DENV-2 | 91.0 (407/447) | 82.3 (28/34) | 93.1 (94/101) | 92.6 (100/108) | 88.3 (91/103) | 92.3 (84/91) |
|  | DENV-3 | 85.5 (382/447) | 82.3 (28/34) | 86.1 (87/101) | 87.0 (94/108) | 84.5 (87/103) | 83.5 (76/91) |
|  | DENV-4 | 84.8 (379/447) | 64.7 (22/34) | 87.1 (88/101) | 88.0 (95/108) | 84.5 (87/103) | 84.6 (77/91) |
| Spondweni Group | ZIKV | 52.6 (235/447) | 32.3 (11/34) | 48.5 (49/101) | 55.5 (60/108) | 54.4 (56/103) | 59.3 (54/91) |
| Japanese Encephalitis Virus Group | WNV | 81.9 (366/447) | 55.9 (19/34) | 84.2 (85/101) | 88.0 (95/108) | 78.6 (81/103) | 84.6 (77/91) |
|  | SLEV | 86.3 (386/447) | 64.7 (22/34) | 87.1 (88/101) | 89.8 (97/108) | 84.5 (87/103) | 90.1 (82/91) |
| Ntaya Virus Group | ILHV | 85.7 (383/447) | 64.7 (22/34) | 85.1 (86/101) | 90.7 (98/108) | 83.5 (86/103) | 89.0 (81/91) |
|  | ROCV | 75.8 (339/447) | 70.6 (24/34) | 74.3 (75/101) | 72.2 (78/108) | 79.6 (82/103) | 78.0 (71/91) |
| Negatives |  | 3.4 (15/447) | 11.8 (4/34) | 4.0 (4/101) | 0.9 (1/108) | 3.9 (4/103) | 2.2 (2/91) |

### Cross-reactivity between principal endemic and emerging Flavivirus es

We performed HIA, a cell-based assay, to assess overall cross-reactivity by performing serial dilution of plasma samples; results are summarized in Table 3 and Figure 2. YFV is endemic in the Amazon region and vaccination is compulsory, here we observed that 92.8% of individuals tested have anti-YFV-17D reactive antibodies compared to 80.5% towards the wild-type YFV (Table 3). Whereas a total of 80.3% self-reported vaccinated for YFV (Table 2). 37.2% self-reported previous DENV infection, whereas we detected 93.7% of individuals with anti-DENV antibodies against at least one serotype. A chi-square test found a significant difference between the observed (laboratory confirmed) and expected (self-reported) dengue infection (p<0.0001, χ2 test, data not shown). Among the DENV serotypes, DENV-2 followed by DENV-1, DENV-3 and DENV-4 had the highest seropositivity (Figure 2 and Table 3). Individuals with high DENV titers correspondingly had higher reactivity to other Flavivirus es (Figure 2A). All samples were collected before or during the ZIKV epidemic in Manaus and we observed that ZIKV, among all viruses tested, had the lowest seropositivity percentage (Figure 2B). The percentage of individuals reactive towards Flaviviruses classified into serogroups is presented in Table 3. Overall, antibody reactivity was the lowest among the 18-19 age group and increased rapidly with age for all arboviruses tested (Table 3, Figure 2, and supplementary Figure 2).

**Figure 2.**
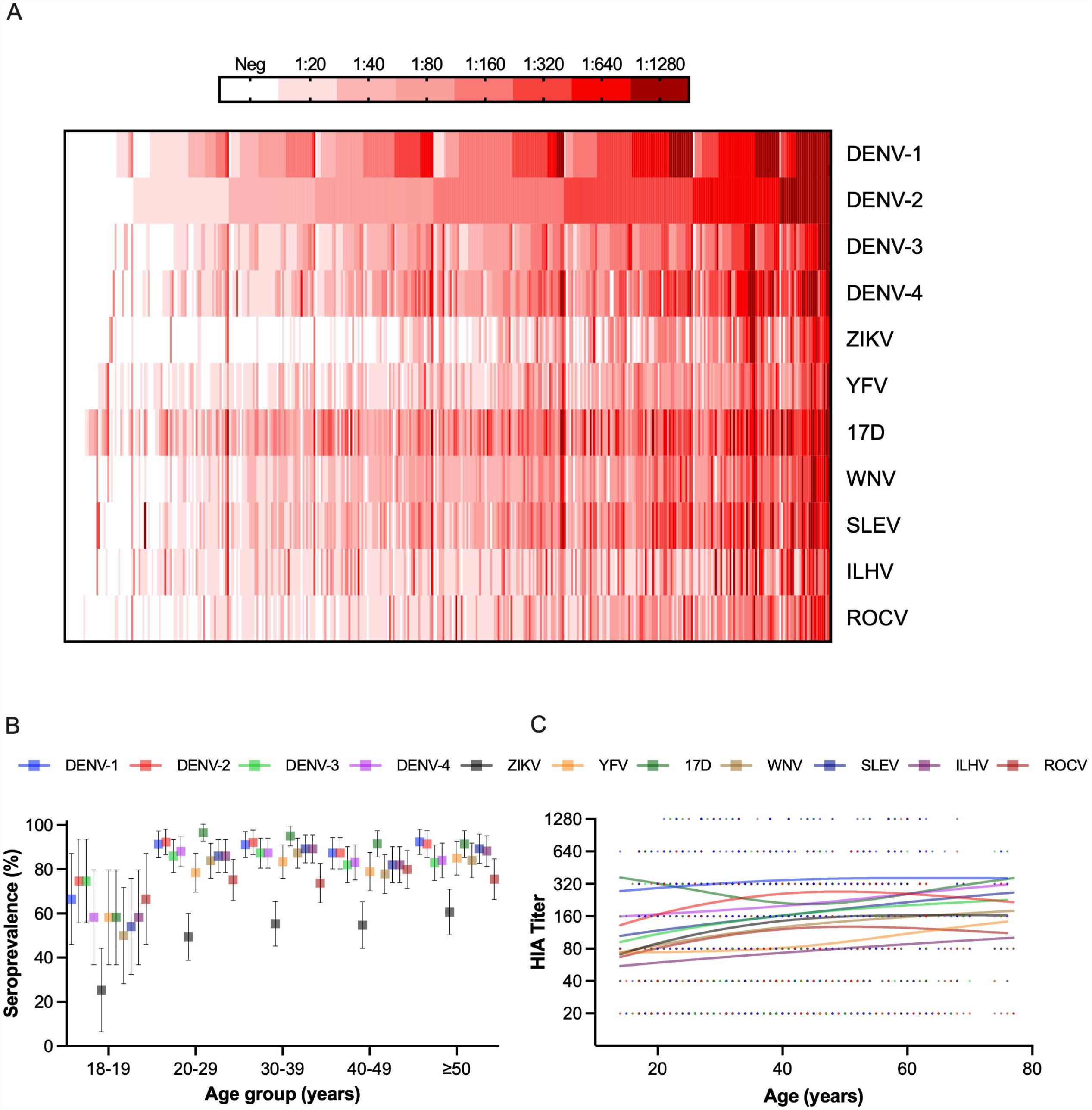
Antibody reactivity profile against emerging and re-emerging human Flaviviruses. Samples (n=447) were tested for antibodies against ten different endemic and emerging Flavivirus es and yellow fever vaccine strain using the Hemagglutination inhibition assay (HIA). (A) Colors depict the HIA titers, each column represents one patient, and each row is one test virus on the heat map. Samples described in the heatmap were sorted as per DENV-2 HIA titers. (B) Samples with HIA titer ≥ 20 units were considered positive and plotted as percentage positives among the study age groups. (C) HIA titers were compared to participant age. Solid lines are splines for each test virus.

Low cross-reactivity between preexisting anti-DENV antibodies and ZIKV Next, we evaluated cross-reactivity to ZIKV among DENV positive individuals using HIA, ELISA, and live virus neutralization assay. First, modified bubble plots compared DENV-1, DENV-2 or ZIKV HIA titers and their distribution for each virus compared to the reference virus. Upon comparing HIA titers, we observed that most of the DENV positive individuals with low titers failed to react with ZIKV. Whereas only individuals with elevated titers for DENV-1 or DENV-2 reacted with ZIKV in the HIA assay (Figure 3).

**Figure 3.**
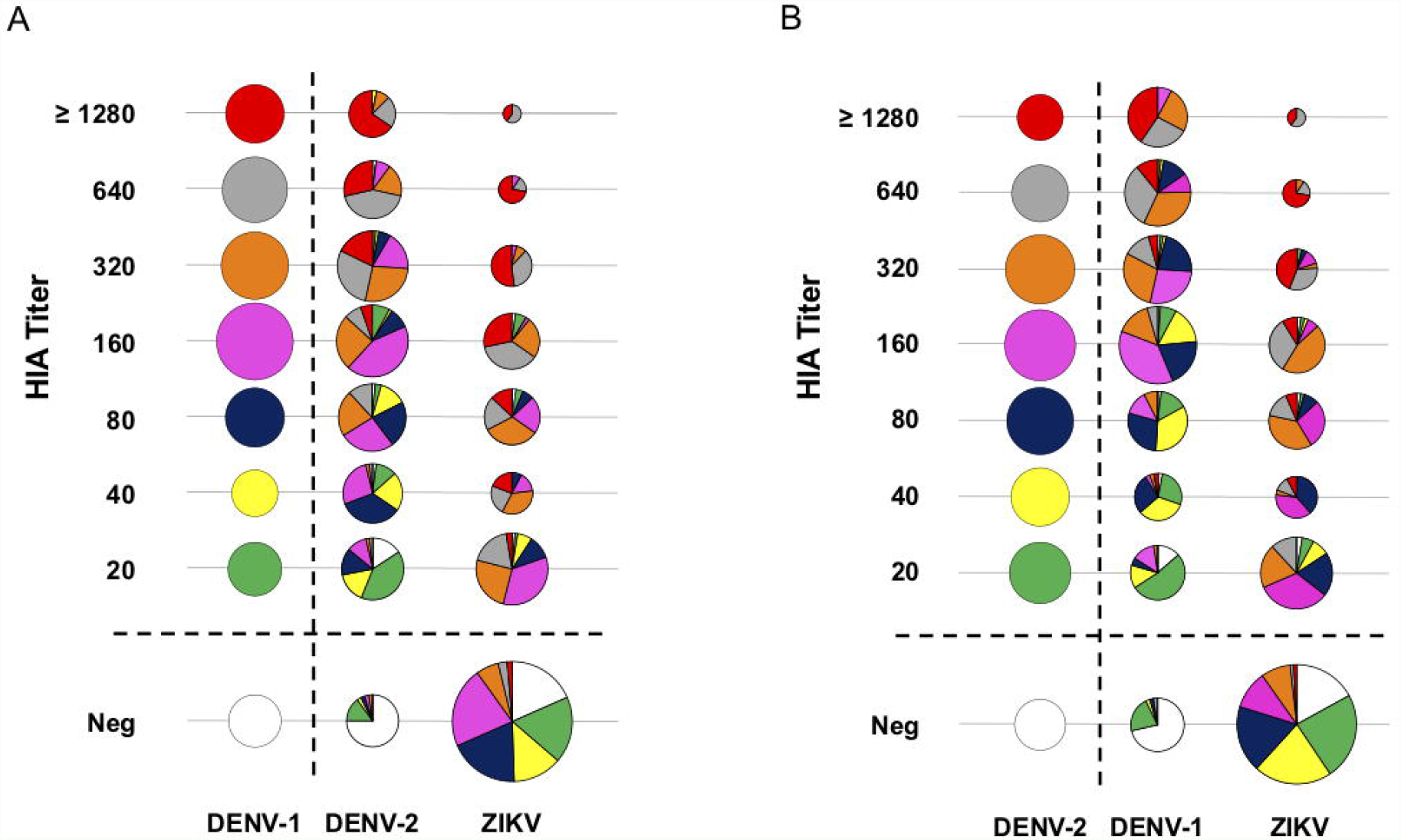
Low cross-reactivity between pre-existing anti-Dengue antibodies and Zika virus. Modified bubble charts compare cross-reactivity (n=447), (A) DENV-1 with DENV-2 or ZIKV, and (B) DENV-2 with DENV-1 or ZIKV. All samples with HIA titer ≥ 20 units were considered positive. Bubble size is proportional to the percentage of individuals with the corresponding HIA titer, and each column adds up to 100%. HIA titers are color-coded for the first reference virus column for comparison. Each pie chart represents the composition of HIA titer for the reference test virus.

Envelope domain III (E-DIII) is an important target for neutralizing antibodies among Flavivirus es, hence we performed an ELISA using E-DIII proteins for DENV-2 or ZIKV and assessed antibody reactivities. Similarly, to HIA, we observed anti-DENV2 E-DIII IgG levels were elevated compared to anti-ZIKV E-DIII IgG antibodies (Figure 4A). A paired analysis of the samples showed a significant decrease in the antibody reactivities between DENV and ZIKV (Figure 4B-C). As expected, anti-DENV E-DIII antibody levels estimated by ELISA significantly increased with age (Figure 4D); and stratified by sex, males showed the highest reactivity towards DENV-2 EDIII (Figure 4E). Splines depict the relationship between HIA antibody titers and ELISA reactivity (Figure 4F).

**Figure 4.**
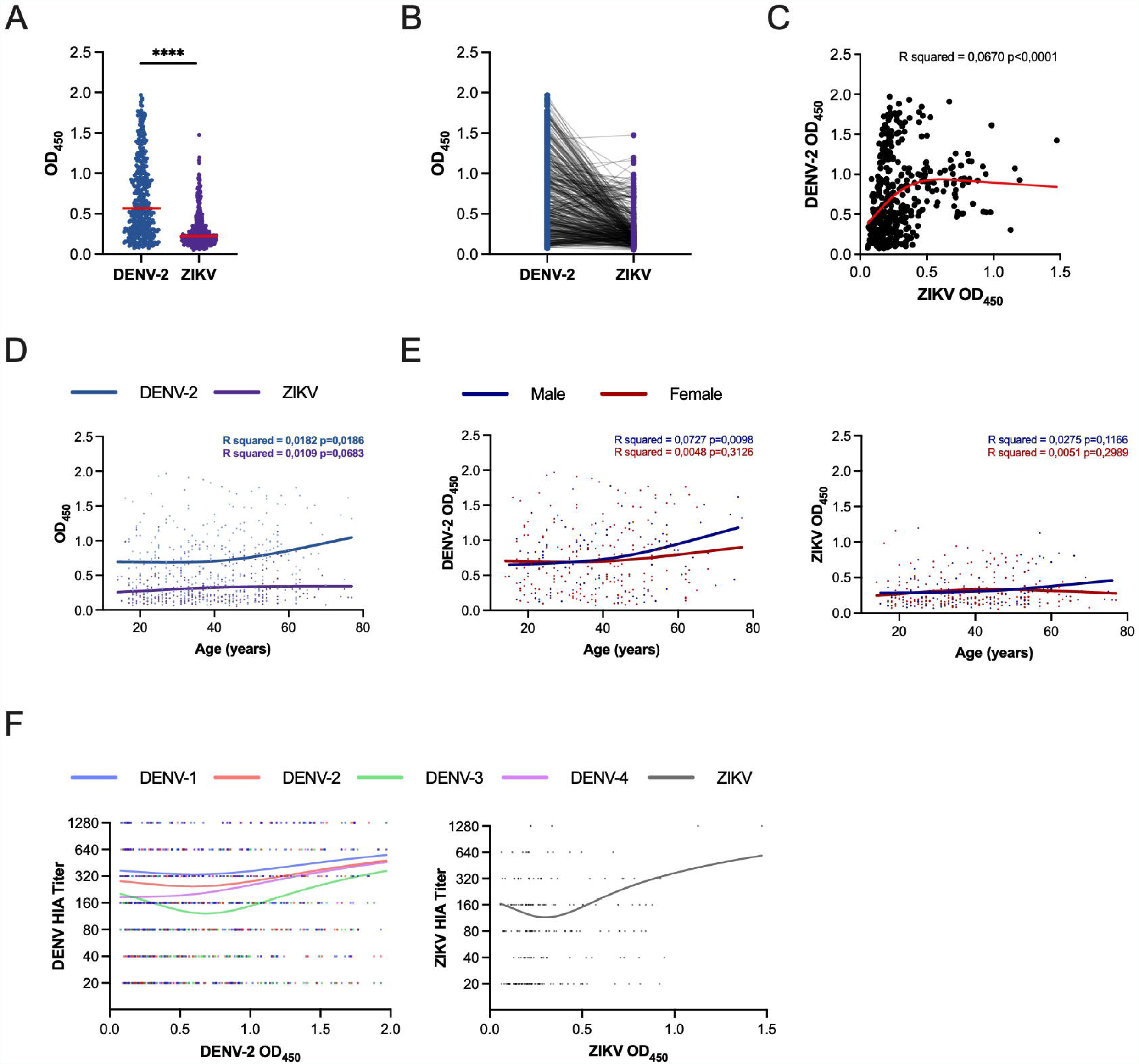
Reactivity between Dengue virus and Zika virus envelope domain III proteins. ELISA plates were coated with DENV-2 or ZIKV envelope domain III (E-DIII) proteins to detect IgG antibodies in human plasma samples (n=416). (A) Range of antibody reactivity towards EDIII antigens is depicted. The red horizontal lines denote median optical density (OD, 450nm) values. (B) Pairwise comparison denotes the reactivity towards DENV-2 and ZIKV. Mann Whitney or paired T test, ****p <0.0001. (C) DENV-2 and ZIKV anti-E-DIII antibody levels measured by ELISA were compared, red line in the graph represents the spline fitting for the distribution. (D-E) DENV-2 or ZIKV E-DIII reactivity was compared to participant (D) age or (E) sex. Solid lines represent splines fit for the antibody distribution. Pearson’s correlation and p-values are denoted in the graphs. (F) HIA tiers for dengue virus serotypes are compared between DENV-2 or ZIKV E-DIII reactivity determined by ELISA. Each solid line is color coded and represents one test virus. Each dot represents one sample in all the graphs.

Furthermore, to understand the DENV antibody reactivity and its relationship to a possible neutralizing antibody response to ZIKV, we performed a Focus Reduction Neutralization Test (FRNT) with live ZIKV in a subset (n=57) of DENV positive samples chosen at random. Figure 5A compares DENV-2 or ZIKV ED-III ELISA results with ZIKV FRNT50 values; samples were divided as higher or lower than the median of reactivity toward the DENV-2 ED-III antigen. Primarily, we observed that individuals with anti-DENV E-DIII OD > Median value (red) neutralized ZIKV more efficiently compared to individuals with OD < median values (green). Percentage relative infection of ZIKV was used to calculate neutralizing antibody titers, each patient is represented by one curve (Figure 5B-D). Overall, DENV-2 E-DIII antibody levels are inversely proportional to ZIKV FRNT values, and individuals with high DENV reactive antibodies had potential neutralizing activity against ZIKV. Collectively, these results indicate that both the quantity and quality of DENV specific response are necessary to neutralize ZIKV.

**Figure 5.**
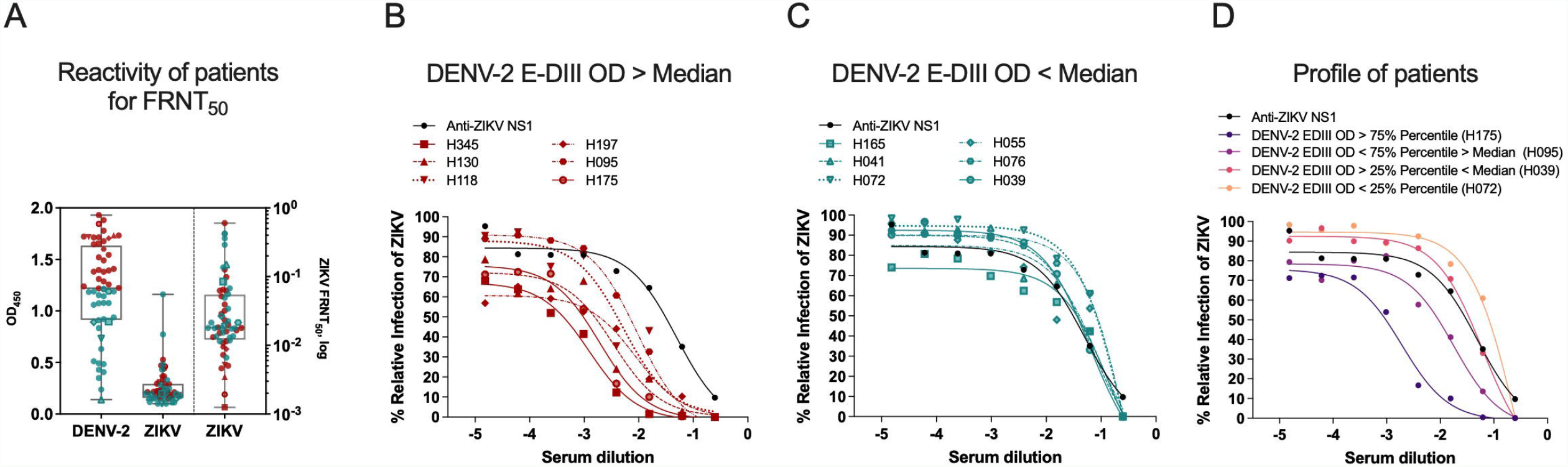
High titers of Dengue virus reactive antibodies neutralize Zika virus. Focus Reduction Neutralization Test (FRNT) for Zika virus was performed to estimate neutralizing antibodies in patient plasma samples (n=57). (A) DENV-2 or ZIKV E-DIII protein specific IgG reactivities are depicted in first two columns. Box and whiskers indicate the median lines, 25 and 75-interquartile and min-to-max points. Red dots denote samples with anti-DENV-2 E-DIII reactivity values greater than the median value and green dots denote samples displaying anti-DENV-2 reactivity less than the median value. Third column in the graph represents the 50-percent FRNT (FRNT50) values for Zika virus. Right Y-axis is FRNT50 values in log for Zika virus. Zika virus antibody neutralization dose-response curves were plotted with (B) high (OD450> median) or (C) low (OD450< median) DENV-2 E-DIII antibody reactivity. Each curve represents one patient sample diluted serially. Anti-ZIKV NS1 monoclonal antibody was used as control. (D) Representative neutralization doseresponse curves were plotted based on DENV-2 E-DIII antibody reactivity.

These findings reinforce the elevated dengue serologic prevalence in the Amazon region and cross-reactivity between Flavivirus es. *Flavivirus* is a genus with highly related species. Since Manaus is endemic to dengue, the presence of high mosquito density may have influenced the zika virus epidemic (Supplementary Figure 1). Here we observed low protection in DENV positive cases with ZIKV. Our results suggest that individuals with high neutralizing titers against DENV could partially neutralize ZIKV. Understanding of cross-reactivity among Flaviviruses is important to comprehend the ZIKV epidemic and for future Flavivirus vaccination and disease control strategies.

## Discussion

Endemic and emerging arboviruses are a huge public health concern worldwide not only due to their unexpected clinical manifestations and potential complications but also due to the lack of appropriate diagnostic assays and vaccines [20-22]. In this study, we report one of the highest dengue seropositivity in Manaus-Brazil [18]; 74.52% by POC and 93.7% by HIA. This confirms that most residents of the region have likely been infected with DENV, if not with other arboviruses at least once. Evidence of varying degree of antibody cross-reactivity with principal emerging Flavivirus es depends on the virus serogroup and preexisting level of antibody titers. Thus, individuals with the highest anti-DENV antibody titers could cross-neutralize ZIKV more efficiently compared to individuals with low anti-DENV titers, which failed to neutralize ZIKV. Elevated dengue prevalence in Manaus and the Amazon region does not undermine the threat to other endemic and emerging arboviruses.

Dengue is endemic in Brazil for at least 40 years and without adequate prevention and treatment, the number of reported cases of dengue has increased in Brazil and other countries of the Americas [23, 24]. In our previous study, we observed an increase in seroprevalence in several Brazilian cities in the last decades [18]. On the contrary, dengue incidence levels have been the same over the last decade in the most regions; nevertheless, there has been a huge increase in severe dengue disease cases and deaths [23-26]. Three out of four Brazilian municipalities are heavily infested with the mosquito *Aedes aegypti*, which highlights the staggering epidemiological and economic burden in the endemic regions [20, 27]. Thus, the prospects of controlling dengue or other arbovirus diseases are not promising. All four DENV serotypes have been detected in Manaus, with the late introduction of DENV-3 and DENV-4 [18, 28, 29]. Our serological results demonstrate a lower seropositivity towards DENV-3 and DENV-4 when compared to DENV-2 and DENV-1. The high DENV prevalence in urban Manaus observed in this study is in line with our previous observations from other Brazilian cities [18]. 37.2% of individuals self-reported a previous DENV infection compared to over 75% individuals with detectable antibodies in this study. This disparity between the observed and expected numbers highlights the need for more laboratory diagnosis of febrile cases.

Yellow fever vaccination is compulsory in Manaus and the Amazon region and 92.8% of the study participants had anti-YFV/17D reactive antibodies, whereas 80.5% had anti-YFV reactive antibodies against the wild-type virus. ILHV and ROCV had been endemic in Brazil, however, their spread in the Amazon region and prevalence studies are lacking. In this present study, the Ntaya virus group represented by ILHV and ROCV had a seropositivity rate of 85.7% and 75.8%, respectively. Recently, WNV [30, 31] and SLEV [32, 33] cases have been reported among horses in Brazil, but no human outbreaks cases have been reported. Cocirculation of these arboviruses that share B and T cell epitopes preludes the understanding of the role of preexisting immunity essential to assess the clinical implications of the disease. Infection with DENV can protect against subsequent JEV, SLEV and WNV infections [34-36], ZIKV infection confers protection against subsequent WNV infection [37], JEV and SLEV infections can protect against WNV and DENV infections [38-41], and ILHV and SLEV can elicit cross-protection against a lethal ROCV challenge [42]. Compulsory YFV vaccination and high prevalence of DENV in Manaus may partially explain the lack of evidence for cocirculation of other Flavivirus es. This cross-protection may partly also explain why YFV is not present in India and other Asian countries, which are endemic to WNV, JEV and DENV, and other Flaviviruses [43]. On the other hand, recent ZIKV outbreak in India demonstrates that cross-protection might be impermanent, and viruses may adapt to epidemiological conditions [44].

The samples used in this study were collected before or during the zika virus epidemic in Brazil. In this present study, 52.6% had anti-ZIKV antibodies, the lowest seropositivity rate among all the viruses tested by HIA. Also, E-DIII ELISA demonstrated a weak reactivity towards ZIKV among DENV positive individuals. Thus, a substantial proportion of participants showed evidence of exposure to DENV but not ZIKV [45]. Moreover, DENV positive individuals with low HIA titers failed to react with ZIKV and only high-titer DENV plasma samples reacted with ZIKV. The role of previous DENV infection in ADE has been controversial; nevertheless, there is little evidence of severe disease after ZIKV infection in DENV positive individuals [8-17]. Patterns of antibody cross-neutralization suggest that ZIKV lies outside the DENV serocomplex [46]; similarly, we observed that HIA was able to distinguish ZIKV from DENV infections when all viruses were analyzed simultaneously. We observed a lack of durable cross-neutralizing antibody response against ZIKV from DENV positive individuals as previously reported [47]. In this study, patients with the highest anti-DENV antibody titers neutralized ZIKV more efficiently, in line with a previous report demonstrating that individuals with elevated DENV titers due to repeated exposure to dengue virus elicit robust cross-neutralizing antibodies against Zika virus [48].

In Manaus, a high prevalence of DENV serotypes and compulsory YFV vaccination together may provide a strong increase in homologous and heterologous cross-neutralizing antibodies. Zika vaccination in a DENV-experienced individual has been shown to boost preexisting Flavivirus immunity and elicit protective responses against both ZIKV and DENV [49]. On the other hand, Tick borne encephalitis virus (TBE) vaccination in YFV vaccinated individuals caused a significant reduction in the TBE-specific neutralizing antibodies [50]. T-cell responses seem to be robust between Flavivirus es and may, at least in part, explain the cross-protection seen against ZIKV from DENV infection [51]. Nevertheless, the durability of humoral and cellular response needs to be assessed in longitudinal studies to determine the role of the preexisting immunity in protection or pathology.

Due to many positive samples, we were unable to perform statistical analysis to identify epidemiological risk factors. Our convenience sampling oversampled females that impede calculation of true dengue prevalence for Manaus city; however, our results demonstrate that urban Manaus DENV prevalence is high, most of the individuals get infected before attaining thirty-years and male-sex had higher antibody titers. We used DENV2 ED-III protein in ELISA to compare with ZIKV ED-III response and did not test with other dengue serotypes; nevertheless, we believe that a similar relationship will be observed given the HIA results and since DENV and ZIKV belong to distinct serogroups. Although DENV neutralizing serotype-specific antibodies are mainly against E-DIII, there might be a sustainable antibody response against other targets, such as domains I/II of the envelope protein, which were not evaluated in this study. We also did not have history of hospitalization or severe disease caused by dengue among our study participants to correlate with the humoral response.

In conclusion, we observed a low cross-protection among dengue positive individuals to ZIKV. However, further longitudinal studies are needed on ADE and the possible role of cross-reactive anti-DENV antibodies in Flavivirus pathogenesis. ZIKV and other emerging Flavivirus es will become increasingly prevalent in DENV-endemic regions, raising the possibility that preexisting immunity to one virus could modulate the response to a heterologous virus, although whether this would be beneficial or detrimental remains unclear and could vary with the combination of the viruses prevalent in the region. The COVID-19 pandemic has negatively affected mosquito control measures; additionally, financial resources have been diverted toward the pandemic, making control measures more difficult. Presently, Brazil and the Amazonian region face a complex epidemiological scenario characterized by simultaneous circulation of several arboviruses and high mosquito density, hence a functional vector control program and febrile syndrome surveillance are essential to identify unforeseen epidemiological threats.

## Data Availability

All data produced in the present work are contained in the manuscript

## Supplementary Figures

Supplementary Figure 1: Dengue and Zika cases evolution through and post-Zika virus epidemic in Manaus city and Brazil.

Dengue and Zika cases per epidemiological week between 2015-2017 in Manaus (A) and Brazil (B) were obtained from the Ministry of Health (DATASUS - https://datasus.saude.gov.br/informacoes-de-saude-tabnet/)and Health Pan-American Organization (PLISA - https://www3.paho.org/data/index.php/es/), date accessed June 30, 2021.

Supplementary Figure 2: Dengue-reactive antibody distribution in the Amazonian population.

Bubble chart is used to represent HIA dengue positive samples as function of antibody titers and age groups. Bubbles show the column percentages for each age group and their sizes are proportional to their values.All samples were considered positive with HIA titer ≥ 20 units.

Supplementary Figure 3: Correlation matrix between the study variables

## Reference

1. Vasconcelos, P.F.d.C., Yellow fever in Brazil: thoughts and hypotheses on the emergence in previously free areas. Revista de Saúde Pública, 2010. 44(6): p. 1144–1149.

2. Brasil, P., et al., Zika Virus Infection in Pregnant Women in Rio de Janeiro. New England Journal of Medicine, 2016. 375(24): p. 2321–2334.

3. Mlakar, J., et al., Zika Virus Associated with Microcephaly. New England Journal of Medicine, 2016. 374(10): p. 951–958.

4. dos Santos, T., et al., Zika Virus and the Guillain–Barré Syndrome — Case Series from Seven Countries. New England Journal of Medicine, 2016. 375(16): p. 1598–1601.

5. de Oliveira, W.K., et al., Zika Virus Infection and Associated Neurologic Disorders in Brazil. New England Journal of Medicine, 2017. 376(16): p. 1591–1593.

6. Goncalvez, A.P., et al., Monoclonal antibody-mediated enhancement of dengue virus infection in vitro and in vivo and strategies for prevention. Proceedings of the National Academy of Sciences, 2007. 104(22): p. 9422–9427.

7. Grifoni, A., et al., Prior Dengue Virus Exposure Shapes T Cell Immunity to Zika Virus in Humans. Journal of Virology, 2017. 91(24).

8. Bardina, S.V., et al., Enhancement of Zika virus pathogenesis by preexisting antiFlavivirus immunity. Science, 2017. 356(6334): p. 175–180.

9. Zimmerman, M.G., et al., Cross-Reactive Dengue Virus Antibodies Augment Zika Virus Infection of Human Placental Macrophages. Cell Host & Microbe, 2018. 24(5): p. 731-742.e6.

10. McCracken, M.K., et al., Impact of prior Flavivirus immunity on Zika virus infection in rhesus macaques. PLoS Pathogens, 2017. 13(8): p. e1006487.

11. Pantoja, P., et al., Zika virus pathogenesis in rhesus macaques is unaffected by pre-existing immunity to dengue virus. Nature Communications, 2017. 8(1).

12. Kuhn, R.J., et al., Primary infection with dengue or Zika virus does not affect the severity of heterologous secondary infection in macaques. PLOS Pathogens, 2019. 15(8).

13. Terzian, A.C.B., et al., Viral Load and Cytokine Response Profile Does Not Support Antibody-Dependent Enhancement in Dengue-Primed Zika Virus-Infected Patients. Clinical Infectious Diseases, 2017. 65(8): p. 1260–1265.

14. Santiago, G.A., et al., Prior Dengue Virus Infection Is Associated With Increased Viral Load in Patients Infected With Dengue but Not Zika Virus. Open Forum Infectious Diseases, 2019. 6(7).

15. Michlmayr, D., et al., Comprehensive Immunoprofiling of Pediatric Zika Reveals Key Role for Monocytes in the Acute Phase and No Effect of Prior Dengue Virus Infection. Cell Reports, 2020. 31(4).

16. Halai, U.-A., et al., Maternal Zika Virus Disease Severity, Virus Load, Prior Dengue Antibodies, and Their Relationship to Birth Outcomes. Clinical Infectious Diseases, 2017. 65(6): p. 877–883.

17. Damasceno, L., et al., Why Did ZIKV Perinatal Outcomes Differ in Distinct Regions of Brazil? An Exploratory Study of Two Cohorts. Viruses, 2021. 13(5).

18. Salgado, B.B., et al., Prevalence of arbovirus antibodies in young healthy adult population in Brazil. Parasites & Vectors, 2021. 14(1).

19. Amaral, M.P., et al., Homologous prime-boost with Zika virus envelope protein and poly (I:C) induces robust specific humoral and cellular immune responses. Vaccine, 2020. 38(20): p. 3653–3664.

20. Mota, M.T.d.O., et al., Mosquito-transmitted viruses - the great Brazilian challenge. Brazilian journal of microbiology : [publication of the Brazilian Society for Microbiology], 2016. 47 Suppl 1: p. 38–50.

21. Musso, D. and P. Desprès, Serological Diagnosis of Flavivirus -Associated Human Infections. Diagnostics, 2020. 10(5).

22. Pierson, T.C. and M.S. Diamond, The continued threat of emerging Flavivirus es. Nature Microbiology, 2020. 5(6): p. 796–812.

23. Salles, T.S., et al., History, epidemiology and diagnostics of dengue in the American and Brazilian contexts: a review. Parasites & Vectors, 2018. 11(1): p. 19–12.

24. Nunes, P.C.G., et al., 30 years of fatal dengue cases in Brazil: a review. BMC Public Health, 2019. 19(1): p. 13–11.

25. Teixeira, M.G., et al., Epidemiological Trends of Dengue Disease in Brazil (2000–2010): A Systematic Literature Search and Analysis. PLoS Neglected Tropical Diseases, 2013. 7(12): p. e2520.

26. Fares, R.C.G., et al., Epidemiological Scenario of Dengue in Brazil. BioMed Research International, 2015. 2015(5): p. 1–13.

27. Tapia-Conyer, R., et al., Dengue: an escalating public health problem in Latin America. Paediatrics and International Child Health, 2012. 32(1): p. 14–17.

28. de Souza Bastos, M., et al., Simultaneous circulation of all four dengue serotypes in Manaus, State of Amazonas, Brazil in 2011. 2012. 45(3): p. 393–394.

29. Nava, A., et al., The Impact of Global Environmental Changes on Infectious Disease Emergence with a Focus on Risks for Brazil. - PubMed - NCBI. ILAR Journal, 2017. 58(3): p. 393–400.

30. Martins, L.C., et al., First isolation of West Nile virus in Brazil. Memorias do Instituto Oswaldo Cruz, 2019. 114(11): p. 1.

31. Costa, E.A., et al., West Nile virus detection in horses in three Brazilian states. bioRxiv, 2021.

32. Weaver, S.C., et al., Isolation of Saint Louis Encephalitis Virus from a Horse with Neurological Disease in Brazil. PLoS Neglected Tropical Diseases, 2013. 7(11).

33. Silva, J.R., et al., A Saint Louis encephalitis and Rocio virus serosurvey in Brazilian horses. Revista da Sociedade Brasileira de Medicina Tropical, 2014. 47(4): p. 414–417.

34. Sather, G.E. and W.M. Hammon, Protection against St. Louis encephalitis and West Nile arboviruses by previous dengue virus (types 1-4) infection. Proc Soc Exp Biol Med, 1970. 135(2): p. 573–8.

35. Price, W.H. and I.S. Thind, Protection against West Nile virus induced by a previous injection with dengue virus. Am J Epidemiol, 1971. 94(6): p. 596–607.

36. Tarr, G.C. and W.M. Hammon, Cross-protection between group B arboviruses: resistance in mice to Japanese B encephalitis and St. Louis encephalitis viruses induced by Dengue virus immunization. Infect Immun, 1974. 9(5): p. 909–15.

37. Vazquez-Calvo, A., et al., Zika virus infection confers protection against West Nile virus challenge in mice. Emerg Microbes Infect, 2017. 6(9): p. e81.

38. Goverdhan, M.K., et al., Two-way cross-protection between West Nile and Japanese encephalitis viruses in bonnet macaques. Acta Virol, 1992. 36(3): p. 277–83.

39. Tesh, R.B., et al., Immunization with heterologous Flavivirus es protective against fatal West Nile encephalitis. Emerg Infect Dis, 2002. 8(3): p. 245–51.

40. Petrovsky, N., et al., An inactivated cell culture Japanese encephalitis vaccine (JE-ADVAX) formulated with delta inulin adjuvant provides robust heterologous protection against West Nile encephalitis via cross-protective memory B cells and neutralizing antibody. J Virol, 2013. 87(18): p. 10324–33.

41. Li, J., et al., Cross-protection induced by Japanese encephalitis vaccines against different genotypes of Dengue viruses in mice. Sci Rep, 2016. 6: p. 19953.

42. Mossel, E., et al., Ilheus and Saint Louis encephalitis viruses elicit cross-protection against a lethal Rocio virus challenge in mice. Plos One, 2018. 13(6).

43. Monath, T.P. and P.F.C. Vasconcelos, Yellow fever. Journal of Clinical Virology, 2015. 64: p. 160–173.

44. TOI Kerala on alert as zika virus cases rise to 14: Key developements. Times of India, 2021.

45. Hobman, T.C., et al., The possible role of cross-reactive dengue virus antibodies in Zika virus pathogenesis. PLOS Pathogens, 2019. 15(4).

46. Montoya, M., et al., Longitudinal Analysis of Antibody Cross-neutralization Following Zika Virus and Dengue Virus Infection in Asia and the Americas. The Journal of Infectious Diseases, 2018. 218(4): p. 536–545.

47. Collins, M.H., et al., Lack of Durable Cross-Neutralizing Antibodies Against Zika Virus from Dengue Virus Infection. Emerging Infectious Diseases, 2017. 23(5): p. 773–781.

48. Hattakam, S., et al., Repeated exposure to dengue virus elicits robust cross neutralizing antibodies against Zika virus in residents of Northeastern Thailand. Scientific Reports, 2021. 11(1).

49. Dussupt, V., et al., Potent Zika and dengue cross-neutralizing antibodies induced by Zika vaccination in a dengue-experienced donor. Nature Medicine, 2020. 26(2): p. 228–235.

50. Bradt, V., et al., Pre-existing yellow fever immunity impairs and modulates the antibody response to tick-borne encephalitis vaccination. npj Vaccines, 2019. 4(1).

51. Subramaniam, K.S., et al., Two Is Better Than One: Evidence for T-Cell Cross-Protection Between Dengue and Zika and Implications on Vaccine Design. Frontiers in Immunology, 2020. 11.

