## Supplemental Figure 1 for "Antibody cross-reactivity and evidence of susceptibility to emerging Flaviviruses in the dengue-endemic Brazilian Amazon"

Supplementary Figure 1: Dengue and Zika cases evolution through and post-Zika virus epidemic in Manaus city and Brazil.

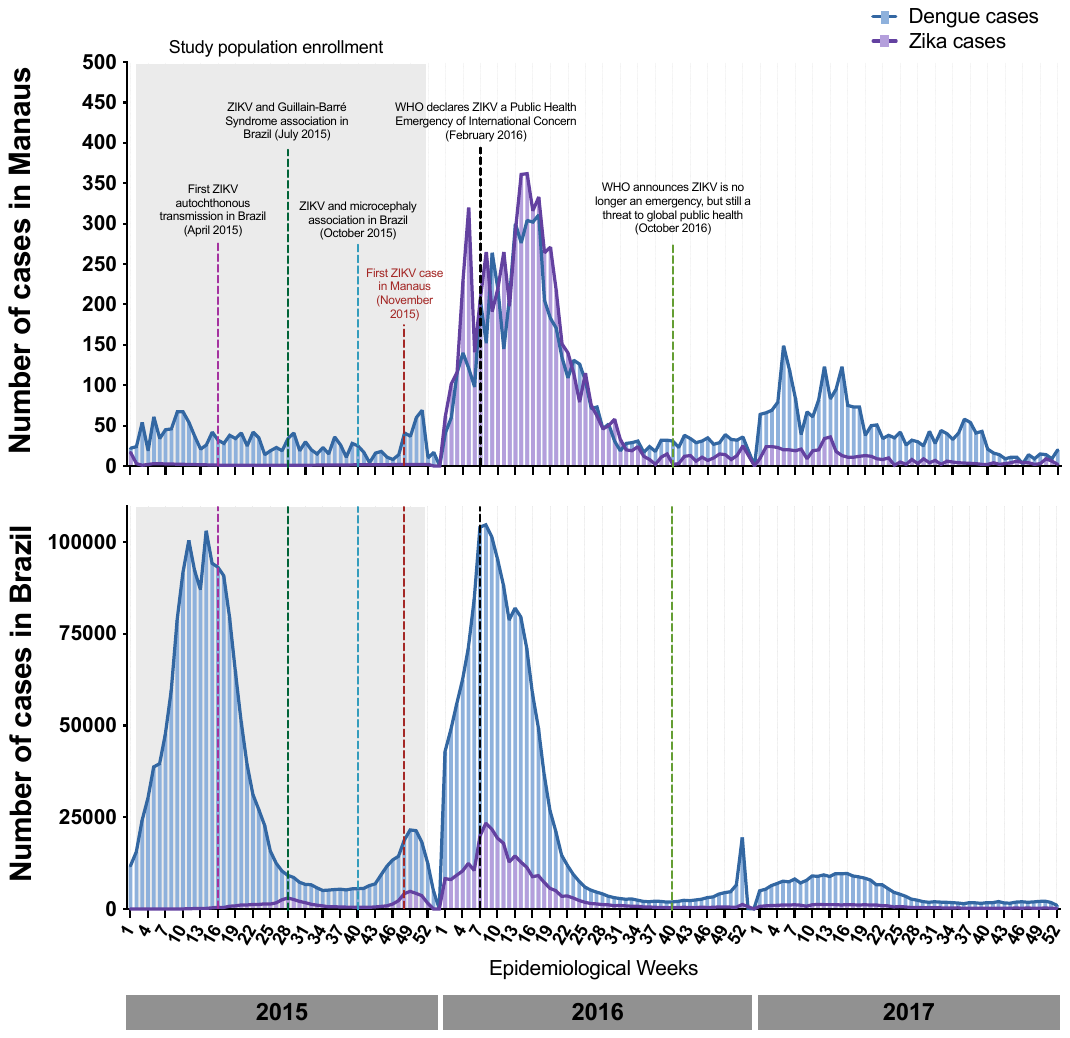

Dengue and Zika cases per epidemiological week between 2015-2017 in Manaus (A) and Brazil (B) were obtained from the Ministry of Health (DATASUS - <https://datasus.saude.gov.br/informacoes-de-saude-tabnet/)>and Health Pan-American Organization (PLISA - https://www3.paho.org/data/index.php/es/), date accessed June 30, 2021.
