## Supplemental Figure 2 for "Antibody cross-reactivity and evidence of susceptibility to emerging Flaviviruses in the dengue-endemic Brazilian Amazon"

Supplementary Figure 2: Dengue-reactive antibody distribution in the Amazonian population.

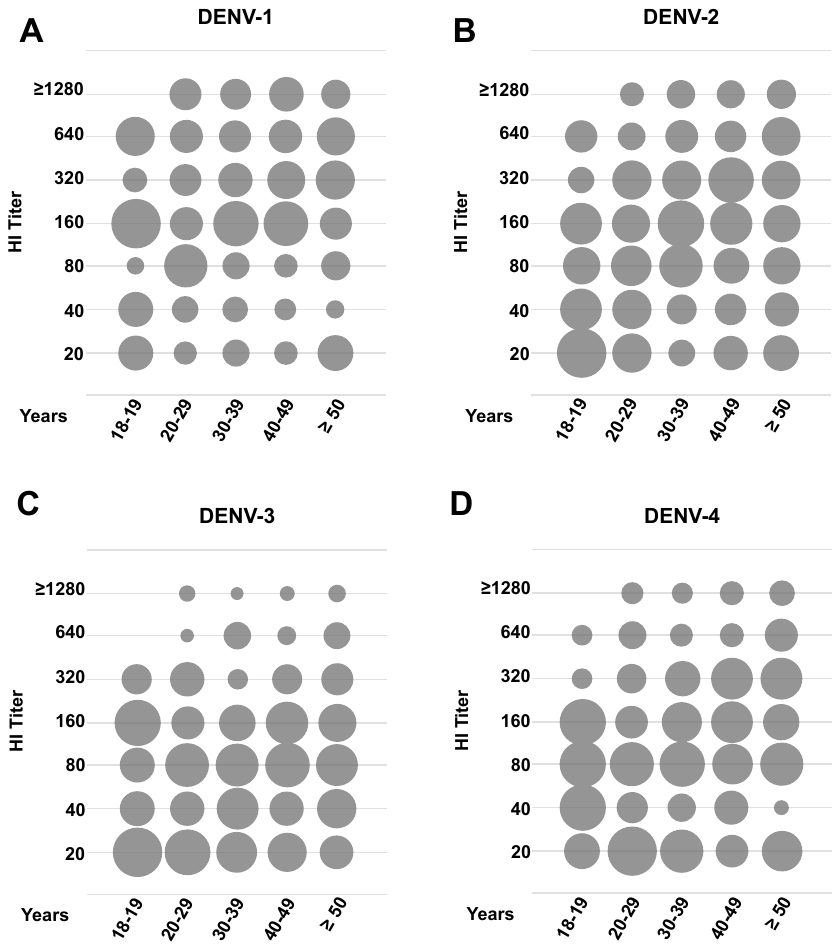

**HIA Titer**

**HIA Titer**

**HIA Titer**

**HIA Titer**

Bubble chart is used to represent HIA dengue positive samples as function of antibody titers and age groups. Bubbles show the column percentages for each age group and their sizes are proportional to their values.All samples were considered positive with HIA titer ≥ 20 units.
