## Supplementary figures and images for "Antibody cross-reactivity and evidence of susceptibility to emerging Flaviviruses in the dengue-endemic Brazilian Amazon"

### Supplemental Figure 3

Supplementary Figure 3: Correlation matrix between the study variables

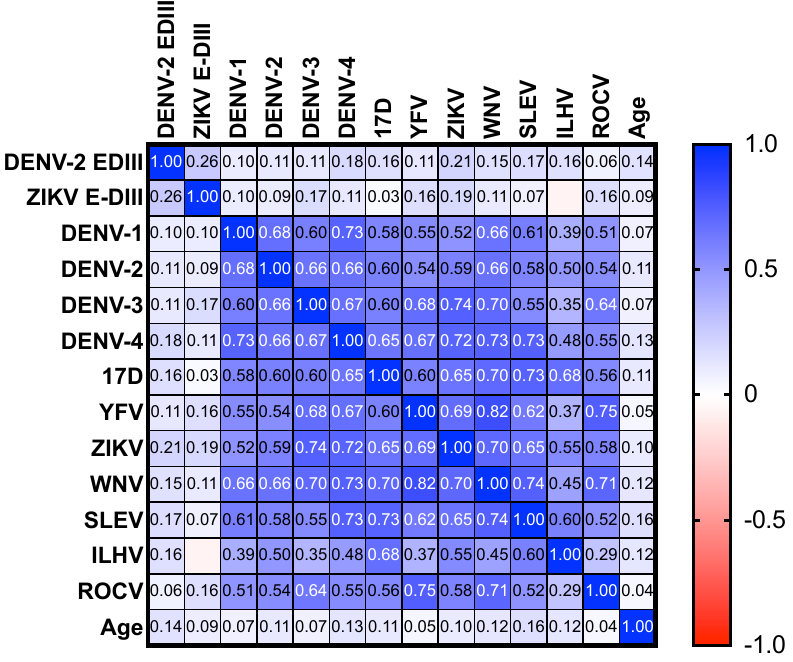
